# Patient-Related Metadata Reported in Sequencing Studies of SARS-CoV-2: Protocol for a Scoping Review and Bibliometric Analysis

**DOI:** 10.1101/2023.07.14.23292681

**Authors:** Karen O’Connor, Davy Weissenbacher, Amir Elyaderani, Ebbing Lautenbach, Matthew Scotch, Graciela Gonzalez-Hernandez

## Abstract

**Background:** There has been an unprecedented effort to sequence the SARS-CoV-2 virus and examine its molecular evolution. This has been facilitated by the availability of publicly accessible databases, the Global Initiative on Sharing All Influenza Data (GISAID) and GenBank, which collectively hold millions of SARS-CoV-2 sequence records. Genomic epidemiology, however, seeks to go beyond phylogenetic analysis by linking genetic information to patient characteristics and disease outcomes, enabling a comprehensive understanding of transmission dynamics and disease impact.

While these repositories include fields reflecting patient-related metadata for a given sequence, inclusion of these demographic and clinical details is scarce. The extent to which patient-related metadata is reported in published sequencing studies and its quality remains largely unexplored.

**Methods:** The NIH’s LitCovid collection will be used for automated classification of articles reporting having deposited SARS-CoV-2 sequences in public repositories, while an independent search will be conducted in PubMed for validation. Data extraction will be conducted using Covidence. The extracted data will be synthesized and summarized to quantify the availability of patient metadata in the published literature of SARS-CoV-2 sequencing studies. For the bibliometric analysis, relevant data points, such as author affiliations and citation metrics will be extracted.

**Discussion:** This scoping review will report on the extent and types of patient-related metadata reported in genomic viral sequencing studies of SARS-CoV-2, identify gaps in this reporting, and make recommendations for improving the quality and consistency of reporting in this area. The bibliometric analysis will uncover trends and patterns in the reporting of patient-related metadata, including differences in reporting based on study types or geographic regions. Co-occurrence networks of author keywords will also be presented. The insights gained from this study may help improve the quality and consistency of reporting patient metadata, enhancing the utility of sequence metadata and facilitating future research on infectious diseases.

**Strengths and Limitations:**

- Classification of papers using machine learning will help pinpoint more relevant papers reducing time to sift through articles for inclusion.
- Study will provide an overview of the availability of information required to facilitate genomic epidemiology studies of SARS-CoV-2
- The bibliometric analysis will analyze publication trends, author affiliations, citation metrics, and other bibliographic information to provide several insights into the broader landscape of research that includes patient metadata reporting in genomic viral sequencing studies of SARS-CoV-2
- As with any search method for relevant studies, some may be missed by both the automated methods proposed and manually developed queries.

## Introduction

Since the onset of the COVID-19 pandemic, there has been an unprecedented effort in genomic epidemiology to sequence the virus, study its transmission, and examine molecular evolution. Public repositories, such as the Global Initiative on Sharing Avian Influenza Data (GISAID)^1^ and NCBI’s GenBank^2^ host millions of SARS-CoV-2 sequence records. As of July 2023, GISAID contains 15.7 million sequences, while 7.7 million have been deposited in GenBank.

The availability of this vast amount of genomic data has facilitated significant discoveries, particularly in phylogenetic and phylodynamic studies ^3–5^. Beyond phylogenetic studies, genomic epidemiology aims to understand the transmission dynamics, evolution, and impact of infectious diseases by analyzing the genetic information of pathogens and linking it to patient demographics and disease outcomes ^6,7^. This work enables the tracking of the spread of pathogens, identifying high-risk populations, and discovering genetic factors that influence disease transmission, severity, and treatment response^6,8^. This knowledge can, in turn, inform public health strategies, guide the development of targeted interventions, and improve the overall understanding of infectious diseases^9^.

Ideally, patient geographic, demographic, and clinical information (such as disease severity and outcome) should be included in the sequence metadata upon its submission to the repository. Both GISAID and GenBank provide the location of the infected host (LOIH) information in their sequence metadata, however, the reported location granularity may vary and often lacks important details such as patient travel history. Similarly, patient demographic and clinical information is rarely complete: approximately 60% of sequences in GISAID have the age or gender of the infected host of the sequence entered as unknown (e.g., ‘Not Available’, ‘Declined’, ‘Not Reported’, etc.) (Figure 1), while GenBank lacks any standardized fields to include this information with sequence submissions.

**Figure 1:**
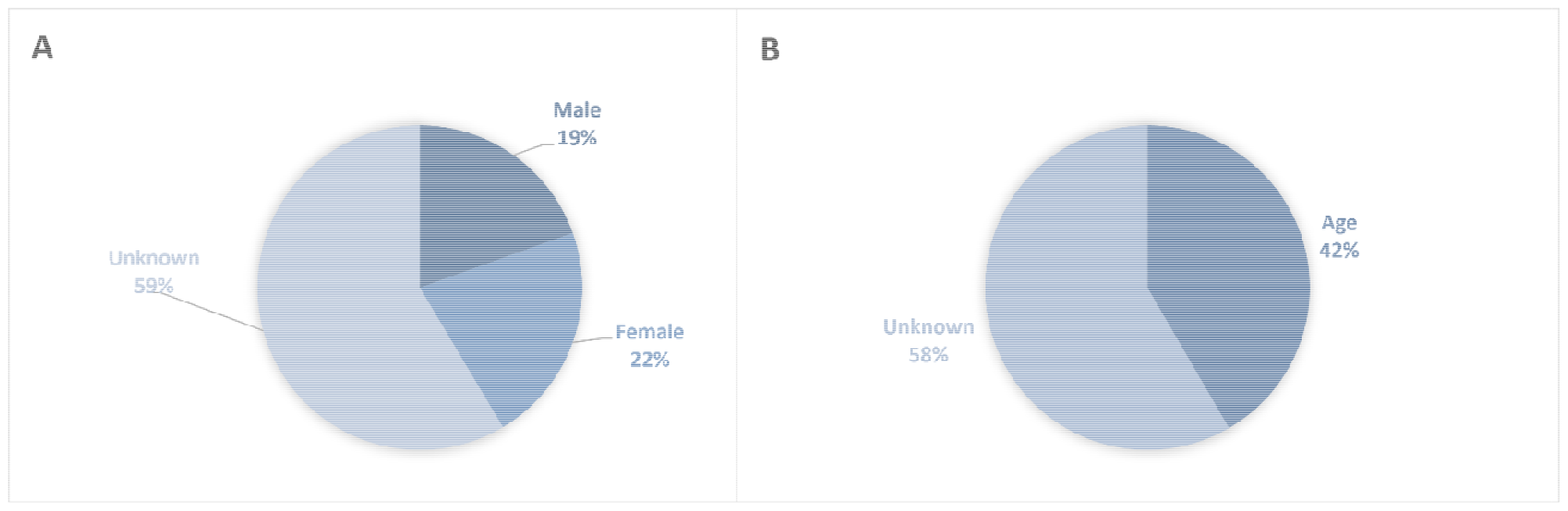
Percent of sequences with reported gender (A) or age (B) in GISAID*. *data downloaded April 3, 2023, representing 15.3 million sequences

This patient information, or at least a subset of it, may be reported in the published studies of those who obtained and performed the genomic sequencing. However, the extent to which location information as well as patient demographic or other clinical information is reported in SARS-CoV-2 sequencing studies remains largely unexplored. Our review aims to bridge this gap in understanding by quantifying the extent and types of patient-related metadata reported in genomic viral sequencing studies of SARS-CoV-2.

Traditionally, identifying studies for a review requires the development of a detailed search strategy of databases using keywords and index terms, querying the title and abstracts of published articles. The selection of keywords greatly influences search results, leading to potential missed or irrelevant studies. Moreover, for the particular focus of our study, discussions of sequencing are often confined to the methods section of papers, rendering title and abstract screening less informative. While a substantial number of research articles related to SARS-C0V-2 and the pandemic have been published, there is sparse linkage between the sequence and publication databases making it difficult to identify publications relevant to the sequences. To overcome these limitations, we propose using an automated classifier to identify relevant studies for review.

A bibliometric analysis uses different methods and data points to quantify the trends and assess the impact of publications in a specific field ^10^. While several bibliometric analyses have investigated COVID-19 related research trends, in general, ^11–13^ and in specific fields such as neurology^14^, long Covid ^15^ and medical imaging ^16^, or specific geographic locations such as Africa^17^, no analysis has specifically focused on the publication trends related to the reporting of patient metadata related to SARS-CoV-2 genomic sequences. Our aims with this review and analysis are to identify reporting and publication trends as well as highlight the gaps in reporting that may hinder the advancement of genomic epidemiology studies of the COVID-19 pandemic.

### Primary Research Objectives

1. To quantitatively assess the extent and quality of patient-reported metadata, including demographic, clinical, and geographic information, in articles reporting original whole genome sequencing of the SARS-CoV-2 virus.
2. To perform a comprehensive bibliometric analysis to ascertain differences and discernible patterns between articles that include patient metadata and those that do not, thereby providing insights into the characteristics and factors associated with the reporting of patient data in the literature.
3. To evaluate the efficacy and reliability of a machine learning classifier in accurately identifying relevant articles for inclusion in the scoping review, enhancing the efficiency and effectiveness of the study selection process.

## Methods

Our review will follow the methodological framework identified by Arksey and O’Malley^18^. The scoping review will be reported in line with the PRISMA-SrC checklist ^19^.

### Data Sources

We will utilize the NIH’s LitCovid collection (1) for our machine learning classification. LitCovid is a curated collection of scholarly articles related to the Coronavirus Disease 2019 (COVID-19). The collection contains over 378,000 publications from 8,000 journals and is updated daily. LitCovid includes published articles as well as preprints. Additionally, we will independently search PubMed directly using a two-faceted search strategy and the NCBI E-utilities program to find publications linked to sequences. This combined approach will help ensure a comprehensive coverage of the literature for our study.

### Search Strategy

#### Classification Model

Our classification model will be trained using manually annotated data. A full-text search strategy was developed to filter the LitCovid collection resulting in a corpus of targeted articles for annotation. The papers identified through the pipeline were annotated by two experienced annotators using the Inception annotation tool ^20^. The annotators reviewed the full text of the articles and labeled sentences which confirmed the study’s performance of SAR-CoV-2 sample sequencing from human specimens. The classifier will be instantiated as a pre-trained neural network, specifically a transformer model called BERT-base-uncased.

#### Search Strategy

To evaluate our classifier and identify studies that may have been missed due to classification errors or lack of full text in the LitCovid collection, we will create a search strategy to independently search PubMed. We will develop a two-faceted search strategy to find “SARS-CoV-2” and “whole genome sequencing” related publications. We will utilize the search strategy developed for the LitCovid collection with additional keywords added to identify studies that report whole genome sequencing. Additionally, we will search for publications linked to SARS-CoV-2 sequences using the NCBIs E-utilities eLink programming API.

A publication date restriction of December 2019 onwards will be used in the searches as this review is focused on SARS-CoV-2 sequencing studies. No language restrictions will be placed on the searches, although financial and logistical restraints will not allow translation from all languages. All results will be uploaded to a Zotero ^21^ library where duplicate results will be removed.

### Inclusion/Exclusion Criteria

Papers positively identified by our classifier and our search results will be reviewed for inclusion in the review based on the criteria outlined in Table 1.

**Table 1.** Inclusion and exclusion criteria for the scoping review.

| Facet | Inclusion Criteria | Exclusion Criteria |
| --- | --- | --- |
| **Sample Origin** | Individual human subject | ☐ Non-human sources (e.g., mice, bats, ferrets)<br>☐ Wastewater<br>☐ Microbiome<br>☐ Cloned/Cell Culture Virus |
| **Sequencing Type** | Whole genomic sequencing | Studies will be excluded if the following sequencing methods were exclusively performed:<br>☐ PCR or LAMP for viral detection<br>☐ Single-cell sequencing<br>☐ Gene expression studies<br>☐ Protocol validation studies on cell culture virus<br>☐ Exome sequencing |
| **Study Design** | Any type of peer-reviewed or preprint study reporting on the original sequencing of SARS-CoV-2 samples. | Any other study design, or a study not per reviewed. |
| **Publication Dates** | December 2019, or later | Before December 2019 |
| **Language** | All | None |

Two reviewers will perform title and abstract screening using the Covidence ^22^ systematic review management tool with any disagreements resolved by discussion. Two independent reviewers will also conduct full-text screening in Covidence.

### Data Extraction

Data extraction will be conducted in Covidence. The reviewers will examine the full text of the articles, including any supplementary files, for data extraction. The customizable interface will be designed to prompt the reviewer to extract various details, such as general publication information, study characteristics, sequencing specifics, and the presence or absence of patient demographic, clinical, or location information. Furthermore, the location of this information within the articles will be noted. An example of the data extraction form can be found in Table 2.

**Table 2:** Example of data that will be extracted from included studies.

| Prompt | Response |
| --- | --- |
| Publication Information |  |
| Study Name | Free text |
| Article Title | Free text |
| Year of Publication | YYYY |
| Publication Type | Journal, Conference, Preprint |
| **Study and Sequence Information** |  |
| Study Objective | Free text |
| Location of Study (country) | Free text |
| Number of patients | Free text |
| Number of samples sequenced | Free text |
| Repository sequences deposited to | GISAID, GenBank, Other, NR |
| For studies with >1 patient, are sequences linked to a patient? | Yes \| No |
| **Patient Demographic Information Reported** |  |
| Age | Yes \| No |
| Gender | Yes \| No |
| Race/Ethnicity | Yes \| No |
| **Patient Clinical Information Reported** |  |
| Symptoms | Yes \| No |
| Severity | Yes \| No |
| Inpatient or Outpatient | Yes \| No |
| Treatments | Yes \| No |
| Outcomes | Yes \| No |
| **Patient Geographic Information Reported** |  |
| Location of Residence | Yes \| No |
| Travel Information | Yes \| No |

We will test the initial extraction form on a subset of articles and revise it as needed.

For bibliometric analysis, all pertinent data points will be extracted for studies included in our review including, author location and institution information, journal, study type, citation metrics, and author keywords.

### Data Analysis

The extracted data will be synthesized and summarized to quantify the availability of patient metadata in the published literature of SARS-CoV-2 sequencing studies using an exported spreadsheet from Covidence. For the bibliometric analysis, data will be analyzed and visualized using the VOSviewer^23^ software or the bibliometrix^24^ package for R. Other software tools may be used as needed for analysis.

## Results

We will summarize and narratively describe our findings, using tables, graphs, and charts when applicable regarding the number of sequences covered in our included studies, the distribution of the sequences in the respective repositories, and the quantity and type of reported patient metadata in the studies. We will also present the geographical location of the study’s authors using maps and report our findings, including the most frequent journals and article types, as well as analyze differences between studies that reported patient data from those that did not. Co-occurrence networks of author keywords will be presented to highlight the frequency and differences in themes and study focus between the reporting groups.

## Discussion

There has been an unprecedented effort in the sequencing and sharing of the viral genomes of SARS-CoV-2 through publicly available databases, such as GISAID and GenBank, during the COVID-19 pandemic. However, the utility of these sequences for genomic epidemiology may not be fully realized due to the unavailability of relevant metadata about the patient from whom the specimen was obtained ^25^. Our study aims to conduct a scoping review and bibliometric analysis focusing on patient metadata reporting in genomic viral sequencing studies of SARS-CoV-2.

This scoping review will provide valuable insights into the current state of reporting of patient-related metadata in SARS-CoV-2 sequencing studies. The review findings will be used to identify gaps in the reporting of patient metadata and make recommendations for improving the quality and consistency of reporting of patient-related metadata in SARS-CoV-2 sequencing studies.

In addition to the findings of our scoping review, the bibliometric analysis will likely identify several other important trends and patterns in the reporting of patient-related metadata. For example, the analysis may find that the reporting of patient-related metadata is more common in certain types of studies, or that it is more likely to be reported in studies from certain geographic regions. The findings of the scoping review and bibliometric analysis will provide valuable insights into the factors that influence the reporting of patient-related metadata and will help to inform future research on this topic. Furthermore, the identification and quantification of the metadata in literature may aid in advancing other research, such as the development of machine learning methods to extract this information and enhance sequence data through automatic methods.

## Strengths/Limitations

This study will conduct a systematic and comprehensive scoping review, encompassing a large number of articles from various databases ensuring a thorough examination of the current state of reporting patient-related metadata in SARS-CoV-2 sequencing studies, and will provide a comprehensive overview of the available literature. The inclusion of bibliometric analysis will go beyond the scoping review to analyze publication trends, author affiliations, citation metrics, and other bibliographic information to provide several insights into the broader landscape of research that includes patient metadata reporting in genomic viral sequencing studies of SARS-CoV-2.

While some relevant studies may be missed due to search limitations and the classification model, our study’s strength lies in providing valuable insights into the current state of reporting patient-related metadata. Although certain limitations exist, such as potential limitations in reported patient metadata^26,27^ and the focus on SARS-CoV-2 sequencing studies, our findings will contribute to improving the quality and consistency of reporting in genomic epidemiology. Future research can build upon our study to address these gaps and enhance reporting practices in this field.

## Conclusion

This protocol outlines the steps that we will take in our scoping review which will be supported by an automated classifier and bibliometric analysis. We will fill the knowledge gap regarding the extent and types of patient-related metadata reported in genomic viral sequencing studies of SARS-CoV-2 and will provide valuable insights by identifying themes and trends in the published literature. The results of this study may encourage improved and standardized reporting practices which will significantly enhance the utility of sequence metadata and aid in advancing our understanding of the SARS-CoV-2 or any future pandemic.

## Data Availability

All data produced in the present work are contained in the manuscript

## Declarations

### Ethics and Dissemination

This study will analyze and synthesize previously published information. Data sharing is not applicable to this article as no datasets were generated or analysed during the current study. We will submit for publication the completed scoping review and bibliometric analysis. At that time, any extracted data and data used in our analysis will be made available with the publication.

### Authors’ contributions

KO, EL, MS and GGH designed the study. KO was a major contributor in the writing of the manuscript. DW designed the classification methods. KO and AE designed the annotation methods. All authors read, edited and approved the final manuscript.

## Acknowledgements

Not applicable.

